# Enhanced cholera surveillance as a tool for improving vaccination campaign efficiency

**DOI:** 10.1101/2022.11.25.22282776

**Authors:** Hanmeng Xu, Kaiyue Zou, Juan Dent, Kirsten E. Wiens, Espoir Malembaka Bwenge, Lee M. Hampton, Andrew S. Azman, Elizabeth C. Lee

## Abstract

Systematic testing for *Vibrio cholerae O1* is rare, which means that the world’s limited supply of oral cholera vaccines may not be delivered to areas with the highest true cholera burden. We modeled how expanding *V. cholerae* testing affected vaccine impact and cost-effectiveness across different bacteriological confirmation and vaccine targeting assumptions. Systematic testing yielded higher efficiency and cost-effectiveness and slightly fewer averted cases than status quo scenarios targeting suspected cholera. With a 10 per 10,000 incidence rate targeting threshold, testing and status quo scenarios averted 10.3 (95% PI: 8.3-13.0) and 5.6 (95% PI: 4.6-6.7) cases per 1,000 FVPs, respectively. Comparing these scenarios, testing costs increased by $37 (95% PI: 29-52) while vaccination costs reduced by $376 (95% PI: 275-556) per averted case. Introduction of systematic testing into cholera surveillance could improve efficiency and reach of global OCV supply for preventive vaccination.

---

Cholera remains a major public health threat in areas with limited access to safe water and sanitation services. Africa bears a substantial part of the global burden of cholera with an estimated 87 million people living in high incidence districts (i.e., > 1 suspected case per 1,000 people) [1,2]. However, these estimates rely primarily on data from passive clinical surveillance with infrequent laboratory confirmation and may not reflect true cholera burden.

Cholera incidence varies greatly across space and time. The majority of suspected cases reported in Africa during 2010-2016 were from less than 5% of the population [2,3] and 65% of reported outbreaks during 2010-2019 occurred in only four countries [4]. Even in high incidence populations, cholera transmission can span the endemic-epidemic continuum, including locations with year-round transmission and locations with outbreaks recurring every three to five years and no reported cases in interim periods [5]. This heterogeneity challenges national surveillance systems, which may need multiple case definitions and reporting protocols to accommodate different transmission settings. When both cholera epidemiology and surveillance reporting vary widely, targeting disease control measures efficiently can be extremely difficult.

Previous work has shown that targeting cholera control to areas with high historical burden can make substantial improvements to the cost-effectiveness and public health impact of these interventions [6]. Geographic targeting is critical for disease-specific measures like vaccination; only 36 million doses of oral cholera vaccine (OCV) were produced in 2022 [7]. Yet the cholera surveillance programs required to enable such targeting are lacking. While most cholera-affected countries in Africa perform passive clinic-based cholera surveillance, there is substantial variation in case definitions, reporting coverage, data quality, and case detection practices [3,8–10]. Further, systematic laboratory confirmation of suspected cholera cases through culture and PCR testing is challenging due to limited laboratory resources and supply chains. Among suspected cholera outbreaks in Africa from 2010-2019, laboratory testing data were reported in 25% of outbreaks and only 13% reported at least one confirmed cholera case [4]. While rapid diagnostic tests (RDTs) for *V. cholerae* O1/O139 detection are being increasingly adopted for outbreak detection and case screening, their widespread use is relatively new, performance across tests and in different settings is variable, and global standards for their use and interpretation for surveillance are still in their infancy [8,11].

A recent systematic review and meta-analysis estimated that an average 49% of suspected cholera cases were true cholera, but this proportion varied widely across space and time with a range of 2% to 100% [12]. With suspected cases coming from other diarrhea-causing pathogens like enterotoxigenic *E. coli, Cryptosporidium*, and *Shigella* [13–15], the current practice of prioritizing the world’s limited supply of OCV using primarily suspected cholera surveillance could be highly inefficient. Fine-scale OCV targeting supported by improved bacteriological confirmation capacity would substantially increase campaign efficiency and vaccine impact while simultaneously reducing the number of campaign sites and target population sizes.

Focusing on cholera-affected countries in Africa where district-level cholera incidence estimates are available, we build upon an existing modeling framework [6] to explore the potential gains in vaccination campaign impact and efficiency that may be observed with improved *V. cholerae* O1/O139 confirmation capacity.

## Methods

We modeled the impact of vaccination campaigns under different scenarios and calculated the vaccine impact, vaccination campaign efficiency, and cost-effectiveness relative to a scenario with no vaccination. Here, we describe the model input data and parameters, the components of targeting scenarios, the simulation framework, and calculation of public health and cost-effectiveness metrics.

### Model inputs

#### Suspected cholera incidence rates

Previously published gridded estimates of mean suspected cholera incidence rate from 2010-2016 in 35 countries were downscaled from 20 km x 20 km to 5 km x 5 km by assuming that 5 km x 5 km cells had the same rate as the overlapping 20 km x 20 km cell [2]. We assumed that these rates would remain constant from 2022 through 2035 in the absence of modeled vaccination, but the number of cases could change as the population size changed each year. These gridded estimates were then aggregated to an administrative unit scale as the population-weighted mean of each of its grid cells.

We assumed that a variable fraction of suspected cholera case incidence was due to true cholera infections. To simulate the incidence rate of ‘true cholera’ we multiplied the suspected incidence rate in each administrative unit by a positivity proportion (“*V. cholerae* positivity”). *V. cholerae* positivity was drawn randomly for each administrative unit and simulation, but was assumed constant across years and modeling scenarios. We assumed that *V. cholerae* positivity followed a Beta distribution (alpha = 1.562, beta = 1.638) that was fit to 1000 posterior predictive samples of the pooled adjusted *V. cholerae* positivity from a recent meta-analysis [12].

We defined administrative areas according to the Global Administrative area database (GADM) using the R package GADMtools (version 3.9.1) [16]. Hereafter, first-level administrative units are called “provinces’’ and second-level administrative units are called “districts.”

#### Vaccine properties

As in a previous modeling study [6], we assumed that complete vaccination had a direct protective effect of 66% in the first year, which waned to 0% after five years. Indirect vaccine effects were modeled as a relative (multiplicative) reduction in incidence rate for unvaccinated individuals according to the corresponding grid cell’s vaccination coverage. We assumed 68% of individuals 1 years old and above in the targeted administrative unit received two doses of vaccine in the vaccination year and otherwise none, similar to a previous study [6].

#### Population data

Annual country population estimates and projections were from UN World Population Prospects [17]. The spatial distribution of population within a country followed the relative population proportions of the unconstrained 2020 1 km x 1 km WorldPop population raster after it was aggregated to the 5 km x 5 km resolution [18,19].

### Modeling scenarios

We explored the potential impact and efficiency of targeted cholera vaccine use in scenarios that varied by three primary variables: bacteriological confirmation capacity (three settings), incidence rate thresholds (three levels, above which administrative units would be targeted for vaccination), and administrative scale of the vaccination campaign targeting (two scales). In total, we simulated 18 vaccination scenarios to represent all combinations (3 × 3 × 2) and one ‘no vaccination’ scenario (Figure 1).

**Figure 1.**
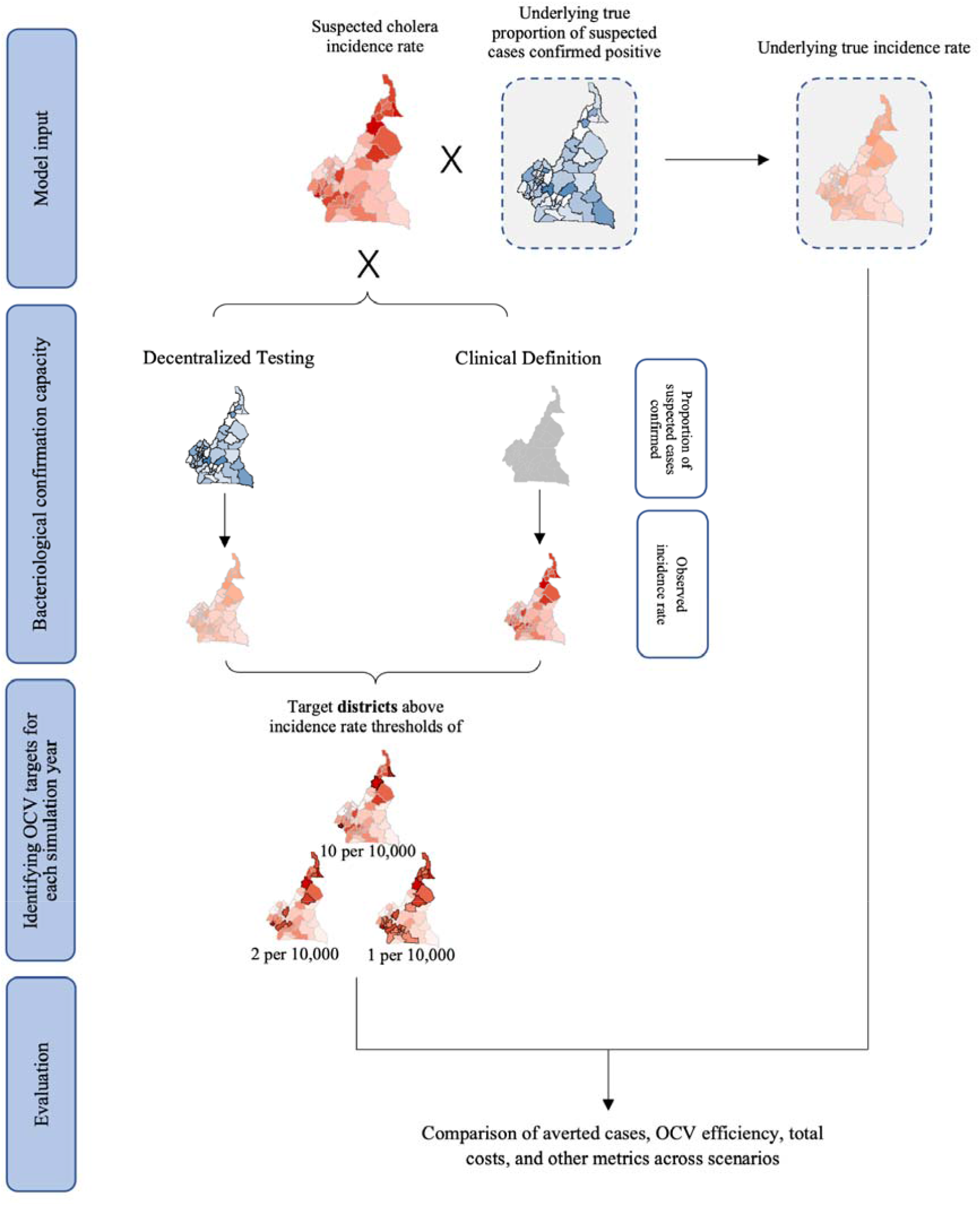
Conceptual depiction of relationship between model inputs and modeling scenarios. Model inputs include suspected cholera incidence rate maps and an underlying true positivity rate, from which a true *V. cholerae* incidence rate map is derived. To determine how OCV is targeted, a bacteriological confirmation capacity setting is applied. Under the Decentralized Testing setting, true positivity rates are assumed to be known at the district-level, and the true incidence rate map is observed. Under the Clinical Definition setting, only suspected cholera incidence is observed. Districts are targeted for OCV in a simulation year if the mean observed incidence rate over the past five years exceeds one of three thresholds, 10 per 10,000, 2 per 10,000, or 1 per 10,000 population, and the location has not been vaccinated in the last 3 years. Models are simulated and public health impact and cost-effectiveness are evaluated with true averted cases, true averted cases per 1,000 fully vaccinated persons (OCV efficiency), and total costs of testing and OCV campaigns, among other metrics.

#### Bacteriological confirmation capacity

Bacteriological confirmation capacity represents a country’s capacity for systematic confirmatory testing. This setting determines how well the incidence rates “observed” by the surveillance system align with true incidence of *V. cholerae*; the observed incidence rate determined where vaccines were allocated.

We considered three bacteriological confirmation capacity scenarios. The low capacity setting, “Clinical Definition”, assumed that only suspected incidence rates were observed as no systematic confirmatory testing was performed (representing the current practice in much of the world). To calculate testing costs, the “Decentralized Testing” setting assumed that suspected cholera samples were tested systematically with RDTs and RDT-positive samples were tested systematically with culture in district-level laboratories, following Global Task Force for Cholera Control (GTFCC) public health surveillance guidance in confirmed outbreak settings (See methodological details in Table S1) [20]. In this setting, the observed and true *V. cholerae* incidence rates were assumed to be the same. The primary manuscript results compared scenarios with “Clinical Definition” and “Decentralized Testing” for district-level campaign targeting.

The “Centralized Testing” setting assumes that suspected cholera samples are tested systematically with culture in a national reference laboratory when calculating testing costs [20], and that these tests experience a 20% reduction in test sensitivity compared to the Decentralized Testing setting; this attenuation is motivated by testing delays and damage to samples that may occur as samples are delivered to a reference laboratory. In this setting, the observed incidence rate was roughly 20% lower than the true *V. cholerae* incidence rate at the national level, with variation derived from district-level *V. cholerae* positivity (See “Suspected cholera incidence rates”).

#### Targeting thresholds

Administrative units were vaccinated if their observed incidence rate exceeded the threshold of 10 cases per 10,000 population, 2 cases per 10,000 population, or 1 case per 10,000 population, according to the scenario.

#### Administrative scale of the vaccination campaign

Vaccination campaign targets were identified at the district or province-level of the country. In scenarios with province-level targeting, the province *V. cholerae* positivity was calculated as the suspected-case-weighted mean of the associated district-level *V. cholerae* positivities.

### Model simulation

We performed simulations independently by country for a duration that enabled three possible rounds of vaccination campaigns (2022-2030) and five additional years for waning vaccine effects (2031-2035). The timeline was chosen to mirror the 2030 cholera control targets set by the 71st World Health Assembly [21]. We modeled all countries in Africa where spatial clinical cholera incidence estimates were available, which included all cholera-affected African countries identified in the GTFCC Global Roadmap to End Cholera [21].

Thirty-five countries were modeled: Angola, Burundi, Benin, Burkina Faso, Central African Republic, Côte d’Ivoire, Cameroon, the Democratic Republic of the Congo, Republic of the Congo, Ethiopia, Ghana, Guinea, Guinea-Bissau, Kenya, Liberia, Madagascar, Mali, Mozambique, Mauritania, Malawi, Namibia, Niger, Nigeria, Rwanda, Senegal, Sierra Leone, Somalia, South Sudan, Chad, Togo, Tanzania, Uganda, South Africa, Zambia, and Zimbabwe.

For scenarios with vaccination, each simulation year proceeds with vaccine targeting and epidemiologic modeling steps during the 2022-2030 vaccination period, and with only epidemiologic steps from 2031-2035 (Table 1). All vaccinated individuals were assumed to be fully vaccinated with two doses. Simulations of the no-vaccination scenario included only the epidemiologic step.

**Table 1.** Description of model simulation steps in vaccination scenarios. The two model components, vaccine targeting and epidemiologic modeling, are implemented in sequential order for each year of vaccination campaigns from 2022-2030. Only the epidemiologic modeling component is implemented in simulation years 2031-2035 to simulate the impact of waning vaccine protection and population turnover after the end of campaigns. A summary of steps within each component is provided.

| Component | Model Simulation Description |
| --- | --- |
| 1) Vaccine targeting | Identify administrative units<sup>1</sup> where the mean observed incidence rate over the past five years exceeds the incidence rate threshold as potential vaccination targets. |
|  | Exclude administrative units that have been vaccinated within the past three years. |
| 2) Epidemiologic modeling | Project the final list of targeted administrative units to grid cell level with the proportion of each cell's population that is vaccinated. |
|  | Calculate the proportion of each gridcell's population that is susceptible to cholera infection based on vaccinated population proportion for the last five years, population turnover, and waning OCV efficacy. |
|  | Model the expected true cases in each grid cell for a given year based on population, susceptible proportion, and true cholera incidence rate rasters, and indirect vaccine protection. |
|  | Calculate observed cholera incidence rate at administrative unit level based on gridded true case model in preparation for vaccine targeting in the following year. |
| <sup>1</sup> Administrative units that do not encompass at least one full 5 km x 5 km grid cell were excluded from the model (Table S4). |  |

### Evaluating public health impact

We calculated true cholera cases averted as the difference between true cholera cases in matched vaccination and no-vaccination simulations. OCV campaign efficiency was defined as the number of true cholera cases averted per 1,000 fully vaccinated persons (FVPs).

Relative efficiency was the ratio of OCV campaign efficiencies in two vaccination scenarios. Percent of FVPs living in high incidence rate administrative units is calculated by dividing FVPs living in units with a true incidence rate that exceeded the scenario’s targeting threshold and the total FVPs in the scenario.

Prediction intervals around these metrics represent stochasticity that was introduced through selection of 200 random posterior draws of mean annual suspected cholera incidence rate maps and random draws from the *V. cholerae* positivity distribution (by district and suspected cholera incidence draw) [12]. A random seed was fixed to enable direct comparison of simulations across modeling scenarios.

### Cost-effectiveness of testing and vaccination

#### Number of tests performed

We followed the GTFCC public health surveillance guidance to determine how many and what kinds of tests were performed under the Decentralized and Centralized Testing settings [20]. The Decentralized Testing setting assumed that the first three suspected cases per health facility per day were tested with RDTs and the first three RDT-positive cases per week per surveillance unit were tested with culture (Table S1). The Centralized Testing setting assumed that the first three suspected cases per health facility per week were tested with culture (Table S1). The percentage of suspected cases tested in the two settings was based on daily clinical surveillance data in four health facilities in endemic cholera transmission regions in the Democratic Republic of the Congo and Bangladesh. We assumed that no tests were performed in the Clinical Definition setting.

#### Cost of testing and vaccination

Based on expert consultation and a brief review of reagent costs from major brands, we assumed that the cost of performing RDT and culture was 1.9 and 13 US dollars (USD) per test, respectively (Table S2, S3) [22]. We assumed that the cost of procuring, shipping, and delivering OCV was 2.36 USD per dose (Table S2) [23–25]. Total costs for a scenario refer to the sum of testing and OCV costs for all tests and doses administered.

#### Evaluating the cost-effectiveness of testing and vaccination

Cost-effectiveness was calculated as total cost per averted case. To understand tradeoffs in testing and vaccination with the introduction of systematic decentralized testing, we subtracted OCV cost per averted case and test cost per averted case between the Clinical Definition and Decentralized Testing scenarios. This subtraction yielded two metrics, OCV cost reduction per averted case and test cost spent per averted case, respectively. OCV cost reduction per test dollar spent is the ratio of OCV cost reduction per averted case and test cost per averted case.

### Data and code access

All modeling and analyses were performed in R version 4.0.3 [26]. The model code, inputs, and explanatory README setup file can be accessed in the Github repository at https://github.com/HopkinsIDD/gavi_vimc_cholera.

## Results

### Systematic testing improves efficiency and targeting of OCV campaigns

Scenarios that targeted OCV campaigns using surveillance with enhanced bacteriological confirmation capacity through systematic testing of suspected cholera cases with RDT and culture (“Decentralized Testing” and “Centralized Testing” scenarios) always had higher OCV efficiency than those that targeted campaigns using only suspected cholera case definitions (“Clinical Definition” scenario) (Table 2, Table S5). When districts with an observed cholera incidence rate over 10 per 10,000 population were targeted with OCV, the Decentralized Testing and Clinical Definition scenarios fully vaccinated 29.1 (95% prediction interval (PI): 21.2-36.5) and 70.9 (95% PI: 65.1-77.8) million individuals and averted 0.3 (95% PI: 0.21-0.38) and 0.4 (95% PI: 0.33-0.47) million cases, thus yielding an OCV efficiency of 10.3 (95% PI: 8.3-13.0) and 5.6 (95% PI: 4.6-6.7) averted cases per 1,000 FVP, respectively. This represented 18.8% (95% PI: 14.9-22.2) of true cases averted in scenarios with systematic decentralized testing and 25.1% (95% PI: 22.4-27.2) in those without.

**Table 2.**
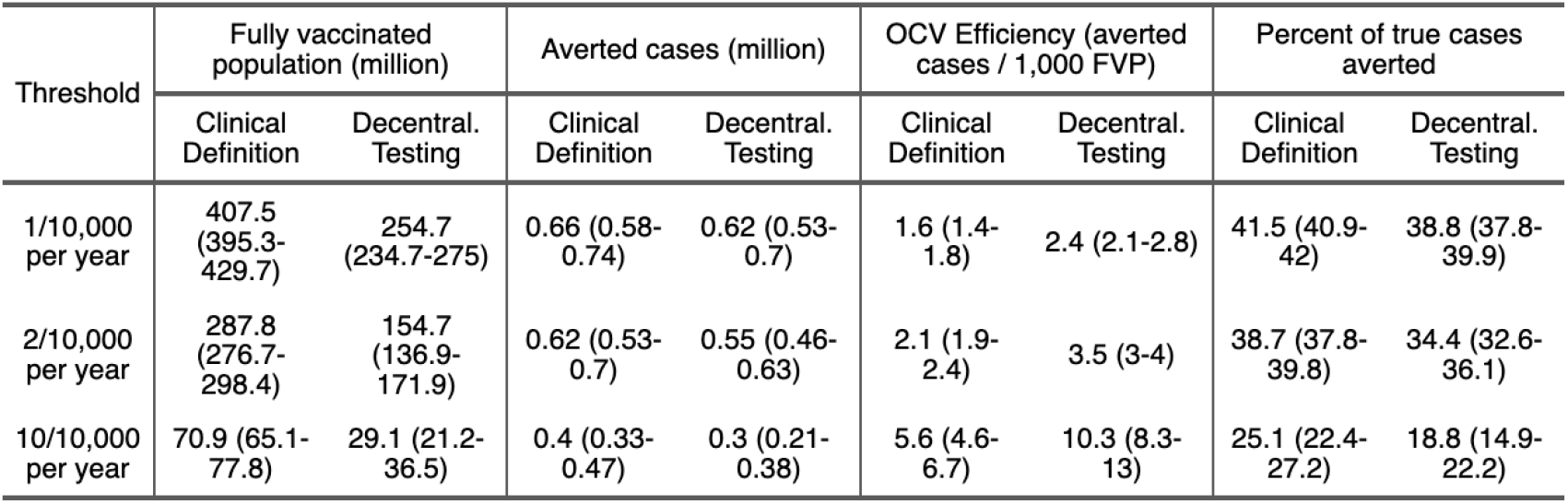
Fully vaccinated persons (FVP), averted cholera cases, OCV campaign efficiency, and percent of true cases averted across different modeling scenarios, 2022-2035. For combinations of modeling scenarios that vary by incidence rate threshold and bacteriological confirmation capacity, we report the median estimates and 95% prediction intervals in parentheses for fully vaccinated persons (FVP), true averted cases (millions), true averted cases per 1,000 FVPs (OCV efficiency), and percentage of true cases averted. “Clinical Definition” refers to a scenario without testing of suspected cases, while “Decentral. Testing” refers to a scenario with systematic testing of suspected cases with RDT and culture in district-level laboratories.

Across targeting thresholds of 1, 2 and 10 cases per 10,000 population, Clinical Definition scenarios vaccinated a median of 1.60 (95% PI: 1.48-1.75), 1.85 (95% PI: 1.67-2.12), and 2.43 (95% PI: 1.98-3.26) times more people than Decentralized Testing scenarios, while averting a median of 1.06 (95% PI: 1.05-1.08), 1.13 (95% PI: 1.10-1.17), and 1.33 (95% PI: 1.22-1.50) times as many true cholera cases, respectively. Thus, OCV efficiency of Decentralized Testing scenarios was 1.50 (95% PI: 1.40-1.63), 1.64 (95% PI: 1.51-1.83) and 1.82 (95% PI: 1.55-2.30) times higher than Clinical Definition scenarios for the three targeting thresholds, respectively (Figure S1).

Scenarios with enhanced confirmation capacity led to a more focused delivery of vaccines to people living in high incidence rate areas. For example, 97.9% (95% PI: 92.9-100%) of FVPs lived in areas where the true incidence rate exceeded 10 per 10,000 population in the relevant Decentralized Testing scenario, in contrast with 40.6% (95% PI: 28.5–49.9%) in the analogous Clinical Definition scenario (Table S5). When targeting areas with an incidence rate above 10 per 10,000, introducing decentralized testing reduced the total number of districts targeted by the OCV campaign from 296 (95% PI: 269-352) to 123 (95% PI: 102-163), and reduced the unique number of districts targeted from 126 (95% PI: 115-147) to 54 (95% PI: 43-71) (Table S5).

### Systematic testing makes OCV campaigns more cost-effective

We found that systematic decentralized testing greatly reduced the combined cost of testing and OCV campaigns across targeting thresholds compared to the Clinical Definition scenario (Table 3). For example, for the targeting threshold of 1 per 10,000, testing reduced the total cost per averted case from 2938 (95% PI: 2569-3329) to 1949 (95% PI: 1712-2287) USD (Table 2). Each USD spent on testing suspected cholera cases reduced OCV campaign costs by 73 (95% PI: 59-86) USD. Decentralized testing also increased OCV efficiency from 1.6 (95% PI: 1.4-1.8) to 2.4 (95% PI: 2.1-2.8) averted cases per 1,000 FVP but averted a slightly lower percent of true cases (38.8%; 95% PI: 37.8-39.9 versus 41.5% (95% PI: 40.9-42) in the Clinical Definition scenario) (Table 2). Increasing the targeting threshold for Clinical Definition scenarios did not achieve the Decentralized Testing scenario outcomes, as testing had the added effect of reducing variability in the detection and targeting of high incidence rate areas (Figure S2).

**Table 3.** Cost-effectiveness of introducing decentralized testing of suspected cholera cases, 2022-2035. We report the median estimates and 95% prediction intervals for total costs per averted cases and metrics that demonstrate tradeoffs in testing and OCV costs between Clinical Definition and Decentralized Testing settings. OCV cost reduction (due to systematic decentralized testing) per averted case is the difference between OCV cost per averted case in the Decentralized Testing and Clinical Definition scenarios. Test cost per averted case is the cost of testing in Decentralized Testing scenarios; no tests were performed in Clinical Definition scenarios. OCV cost reduction per test dollar spent is the ratio of OCV cost reduction per averted case and test cost per averted case.

| Threshold | Total cost per averted case (\$) | | OCV cost reduction per averted case (\$) | Test cost per averted case (\$) | OCV cost reduction per test dollar spent (\$) |
| --- | --- | --- | --- | --- | --- |
|  | Clinical Definition | Decentral. Testing |  |  |  |
| 1/10,000 per year | 2938 (2569-3329) | 1949 (1712-2287) | 978 (757-1247) | 13 (12-15) | 73 (59-86) |
| 2/10,000 per year | 2202 (1953-2533) | 1352 (1181-1581) | 869 (669-1059) | 16 (14-19) | 53 (44-61) |
| 10/10,000 per year | 841 (702-1027) | 494 (396-612) | 376 (275-556) | 37 (29-52) | 10 (8-13) |

### Testing can reduce heterogeneity in OCV cost-effectiveness and lower OCV costs as OCV use expands

Scenarios that introduced decentralized testing into surveillance systems averted slightly fewer true cholera cases than those using a suspected case definition to target OCV (Figure 2A), while vaccinating many fewer people (Figure S3). Consequently, Decentralized Testing scenarios achieved a higher OCV campaign efficiency (Table 2) with lower total costs (Figure 2B). In addition, Decentralized Testing scenarios led to more cost-effective OCV campaigns and reduced across-country heterogeneity in this cost-effectiveness (as measured by OCV cost per case averted) as compared to Clinical Definition scenarios (Figure 2C). Our results suggest that when OCV use expands to include moderate incidence settings, testing becomes more critical to OCV campaign cost reduction (Figure 2D). For example, when we lowered the targeting threshold from 10 per 10,000 to 1 per 10,000 in Nigeria, test costs per true averted case declined from $38 to $15 while the reduction in OCV costs per averted case increased from $367 to $1942 (Figure 2D).

**Figure 2.**
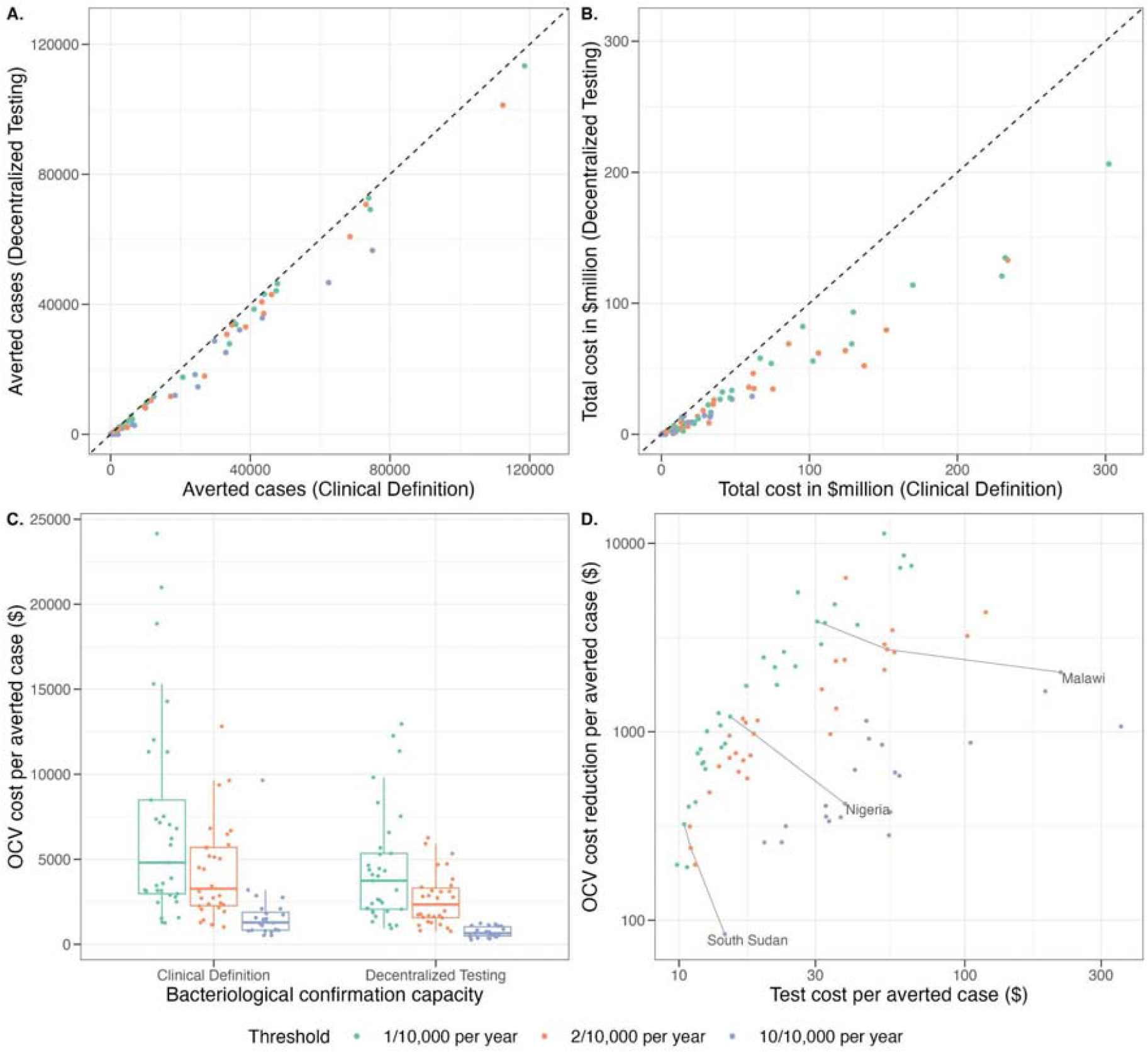
Country-level cost and cost-effectiveness by introducing decentralized testing of suspected cholera cases, 2022-2035. (A) Comparison of averted true cholera cases between “Clinical Definition” and “Decentralized Testing” scenarios. Each point represents a modeled country median across simulations; the x-axis and y-axis are the number of true averted cases in the Clinical Definition and Decentralized Testing scenarios and the dashed line shows where y = x. (B) Comparison of median country total cost (sum of OCV campaign cost and testing cost) between Clinical Definition and Decentralized Testing scenarios. (C) Distribution of median cost of OCV campaign per averted true cholera case by country, under district-level OCV targeting setting. Each point represents the median estimate for one individual modeled country, and the boxplots show the distributions of the country medians. (D) Cost trade-off between decentralized bacteriological confirmation of *V. cholerae* by RDT or culture and OCV campaign, under district-level OCV targeting setting. Each point represents an individual county. The x-axis and y-axis are the median cost spent per averted case and the median cost reduction per averted case, for a specific country. The gray lines link the data points of the three targeting thresholds for three selected countries, South Sudan, Nigeria and Malawi.

### Comparison of effect of decentralized and centralized testing on OCV targeting

We also compared systematic decentralized and centralized testing strategies. Centralized Testing scenarios had slightly higher OCV efficiency and targeted slightly fewer people and administrative units than Decentralized Testing scenarios (Table S5). For example, when targeting districts with an observed incidence rate above 10 per 10,000, the Centralized Testing scenario fully vaccinated 21.6 (95% PI: 15.5-28.9) million individuals, averted 0.23 (95% PI: 0.15-0.31) million cases, and averted 10.8 (95% PI: 8.3-13.3) cases per 1,000 FVPs (Table S5).This represented a more focused campaign with fewer averted cases but greater efficiency than the decentralized testing results reported above, though the 95% prediction intervals overlapped (Table 2). The proportion of FVP living in truly high incidence rate areas was lower in the Centralized Testing scenario, with 79.6% (95% PI: 64.0-92.5%) in the scenario targeting districts above the threshold of 10 per 10,000 (Figure 3, Table S5).

**Figure 3.**
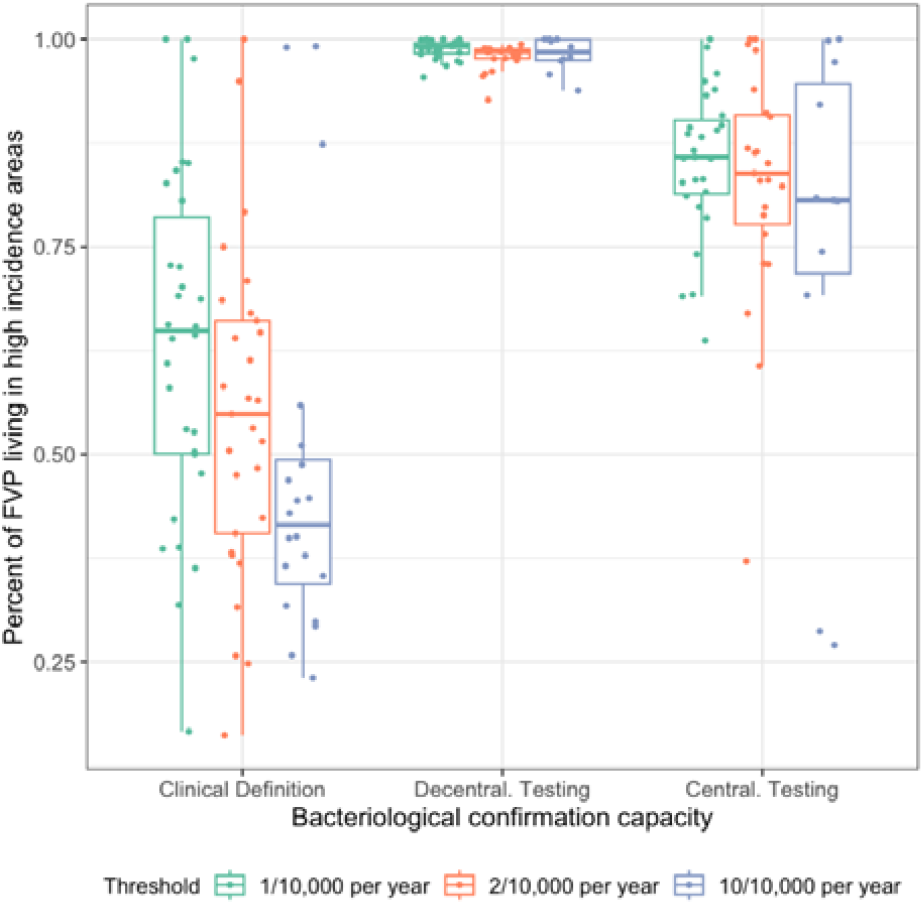
Percent of fully vaccinated persons (FVP) living in high incidence rate administrative units by country. For each modeling scenario, each point represents the country median of percent of FVPs living in administrative units with a true incidence rate exceeding the incidence rate threshold indicated by the color. The boxplot represents the distribution of country-level medians.

## Discussion

Our study builds on previous OCV targeting work [6] by proposing how specific surveillance improvements–such as systematic confirmatory testing of suspected cholera cases–could improve the targeting efficiency of preventive campaigns and extend the reach of the limited global supply of oral cholera vaccines [27]. When districts with an observed mean annual incidence rate over 10 per 10,000 were targeted for vaccination, the introduction of systematic decentralized testing increased OCV campaign efficiency by over 80%, used nearly 84 million fewer doses in a 9-year period, and reduced vaccination costs by $10 for every USD spent on testing, while observing a 25% decrease in true cases averted. In scenarios where OCV use was more expansive (i.e., a lower incidence rate threshold was used to target districts for OCV), the introduction of testing produced smaller gains in OCV campaign efficiency but reduced the number of doses used and the amount spent on testing. This suggests that testing will become even more critical for OCV targeting as preventive OCV use becomes more common in lower burden settings.

Our scenarios compared two systematic testing strategies which were based on recent GTFCC guidance on public health surveillance in areas with confirmed outbreaks [20]. Compared to the centralized testing scenario, which was defined as systematic testing with culture in a single reference laboratory, decentralized testing used a combination of RDT and culture that resulted in a greater total number of tests performed and greater accuracy in the estimated *V. cholerae* positivity rate. The centralized testing scenario had the unanticipated effect of slightly increasing OCV efficiency, as only the highest incidence rate districts remained above the OCV targeting threshold after accounting for reductions in culture sensitivity due to transport and delays. A robust laboratory surveillance system would most likely be a combination of the centralized and decentralized testing scenarios, with perhaps multiple national reference laboratories and smaller, dispersed laboratories with capacity to perform a mix of RDT and culture among suspected cases in their communities [28]. Such a system could gain the advantages from both approaches, with better quality control and standardization of testing in reference laboratories, and improved timeliness in outbreak confirmation from decentralized testing.

Our modeling scenarios do not capture the significant challenges in confirmatory testing faced by many cholera-affected countries. We assumed that reporting and testing of cases would be spatially homogeneous within a district, although differences in access to care, health behavior, health care capacity, and testing capacity could cause spatial heterogeneity. Even variation in antibiotic usage may bias reporting, as this has been observed to reduce culture sensitivity [29]. In addition, our cost-effectiveness analyses do not include the significant resource, training, and supply-chain difficulties of developing and maintaining a laboratory prepared to perform culture at any time.

While several cholera RDTs are now available and beginning to have wide usage, none of the products have received WHO pre-qualification and only recently has Global Task Force on Cholera Control released guidance that integrates RDTs into public health cholera surveillance strategy [11,30,31]. RDT evaluation studies to-date have found high variability in sensitivity and specificity across products and protocols concurrent with concerns about the accuracy of gold-standard culture results [11], leading some to question the best uses of these tools. Nevertheless, use and confidence in cholera RDTs, supported by a comprehensive RDT evaluation with a standardized protocol, is critical for expanding the network of laboratories where decentralized confirmatory testing can be performed.

Our results are limited by the use of mean annual suspected cholera incidence estimates from 2010 to 2016 [2], which depended on care-seeking for cholera symptoms and may not accurately reflect today’s burden. Consequently, it is more appropriate to compare scenarios comprehensively rather than examine the results of any single country. Second, our model projects average burden trends over time, without consideration of high annual variability in cholera transmission (e.g., due to outbreaks, humanitarian emergencies, etc). Finally, we applied a highly simplified approach to vaccine targeting, based only on incidence rates and without vaccine supply constraints.

Robust disease surveillance has long been cited as essential for efficient vaccine targeting and monitoring progress towards disease control [32,33], but previous OCV campaign targets relied primarily on suspected cholera incidence data for prioritization. A recent update to GTFCC guidance now recommends countries to prioritize areas for interventions including vaccination through a multi-dimensional index that includes cholera test positivity from RDTs, culture, or PCR, in addition to suspected case incidence, persistence, and mortality [34]. As confirmed cholera incidence will play an important role in prioritizing targets for future preventive vaccination campaigns, both financial and political investments are needed by ministries of health and the broader global health community to translate diagnostic development into effective surveillance and vaccine distribution for cholera control.

## Data Availability

All data produced in the present study are available upon reasonable request to the authors.

https://github.com/HopkinsIDD/gavi_vimc_cholera

## Acknowledgments

Special thanks to Amanda Debes, Chloe Hutchins, and Allyson Russell for their valuable insights on the costs of *V. cholerae* culture and OCV campaigns.

## Supplementary Material

The supplementary material includes Figures S1 to S3 and Tables S1 to S5.

