## Supplemental Material for "Enhanced cholera surveillance as a tool for improving vaccination campaign efficiency"

**Supplementary Methods	1**

Comparison of Decentralized and Centralized Testing settings	1

Testing and OCV cost parameters	2

Districts excluded from modeling scenarios	3

**Supplementary Results	5**

**References	8**

### Supplementary Methods

#### Comparison of Decentralized and Centralized Testing settings

**Table S1. Different testing strategies, detection sensitivities, and population-level positivity rate values between different bacteriological confirmation capacity settings.**

| **Scenario Setting →**  **Dimension** | **Decentralized Testing** | **Centralized Testing** | **Clinical case detection only & No vaccination scenarios** |
| --- | --- | --- | --- |
| Testing Strategy | Test first 3 suspected cases per day per health facility with RDT (translated to 74% of clinical cases tested by RDT);  Then test 3 RDT-positive cases per week per surveillance unit with culture (translated to 19% of clinical cases tested by culture)  *See notes 1-3* | Test first 3 suspected cases per week per health facility with culture (translated to 30% of clinical cases tested by culture)  *See note 4* | No tests performed |
| Detection Sensitivity | Cholera positivity rate in each district drawn randomly from a distribution with a mean of 49% positivity  *See note 5* | Assume 20% reduction in test sensitivity relative to the district bacteriological confirmation capacity  *See note 6* | Not applicable |
| Population-level Testing Positivity Rate | True confirmed cases are completely observed when targeting OCV. | A pooled, national positivity rate (case-weighted average of the district positivity rates) is used to calculate observed cases; observed cases are used to target OCV. | Not applicable |
| *Testing Strategies were derived from* Public health surveillance for cholera *interim GTFCC guidance for cholera testing in surveillance units with a confirmed cholera outbreak, Table 4* [*[1]*](https://paperpile.com/c/TSkvQI/aNZl)*;*  1) Assume 56% RDTs are positive (unadjusted RDT positivity rate across all settings [[2]](https://paperpile.com/c/TSkvQI/b9ep))  2) Assume 74% of clinical cases are tested with RDT  3) Assume 48% of RDT-positive cases are tested with culture  4) Assume 30% of clinical cases are tested with culture 5) Drawn from a Beta (shape1 = 1.562326, shape2 = 1.638044) distribution (based on Wiens et al., fit a Beta distribution to posterior predictive distribution draws for overall cholera positivity across tests and settings [[2]](https://paperpile.com/c/TSkvQI/b9ep))  6) Multiply cholera positivity distribution by 1-0.2 (20% reduction)[[1]](https://paperpile.com/c/TSkvQI/aNZl) | | | |

#### Testing and OCV cost parameters

**Table S2. Proposed cost parameters for testing and OCV campaign.**

| **Type of Cost** | **Estimate (USD)** | **Data source** |
| --- | --- | --- |
| Cost per Crystal VC Rapid Diagnostic Test (RDT) | 1.9 | UNICEF supply division costs for Crystal VC test kits[[3]](https://paperpile.com/c/TSkvQI/hULn) |
| Cost per culture test | 13 | Assumption informed by expert estimation |
| Procurement cost per OCV in a 1 dose presentation, plastic tube | 1.65 | OCV price in 2023 proposed in UNICEF report[[4]](https://paperpile.com/c/TSkvQI/pYcT) |
| Shipping cost per OCV dose | 0.06 | UNICEF Emergency Stockpile Availability Report, February 2023[[5]](https://paperpile.com/c/TSkvQI/PlyF). |
| Delivery cost in the field per OCV dose | 0.65 | The upper limit of delivery support per dose in USD that a country can apply for from Gavi (through Operational Cost Grants),; a value used for Gavi financial forecasting[[6]](https://paperpile.com/c/TSkvQI/v3NQ). According to Allyson, most countries are actually in the 0.45-0.55 range. |

**Table S3. Estimated cost of *V. cholerae* culture in US dollars.** This table includes rounded, estimated costs of reagents and materials required to perform culture of *V. cholerae* from suspected case stool samples stored in agar stabs which as solicited by expert information. Our model assumption of $13 USD per sample falls within the range presented in the table. The costs listed in [B] are the estimated total costs for a box or bottle of reagent or material. The term "reagent unit" [C-E] refers to the number of preparations that can be made from the amount of reagent or material listed in [A]. Cost per reagent unit [D] is calculated as the Reagent/ Material cost [B] divided by Reagent units available [C]. In some cases, multiple preparations may be required to test a single sample [E]. Lower and upper estimated costs per sample [F-G] are calculated as the product of Cost per reagent unit [C] and Reagent units used per sample [E]. These estimates do not take into account the costs of plastic consumables or running a laboratory.

| Reagent/ Material  [A] | Reagent/ Material cost [B] | Reagent units available [C] | Cost per reagent unit [D] | Reagent units used per sample [E] | Lower est. cost per sample  [F] | Upper est. cost per sample  [G] |
| --- | --- | --- | --- | --- | --- | --- |
| TCBS  (500 g) | 100~320 | 568 | 0.18~0.56 per 10 ml plate | 2~6 plates | 0.35 | 3.38 |
| TSB  (500 g) | 60~200 | 1667 | 0.04~0.12 per 10 ml plate | 3~4 plates | 0.11 | 0.48 |
| Agar powder (500 g) | 160~500 | 3846 | 0.04 per 10 ml plate | 3~4 plates | 0.12 | 0.52 |
| Filter paper (1000 filters) | 120 | 1000 | 0.12 per disk | 4 disks | 0.48 | 0.48 |
| APW (500 g) | 105~180 | 1667 | 0.06 per 5 ml aliquot | 1~2 5 ml aliquots | 0.06 | 0.22 |
| Oxidase  (50 pieces) | 105 | 50 | 2.10 per piece | 1 piece | 2.10 | 2.10 |
| Antiserum - Polyvalent  (2 ml) | 165 | 2 | 82.50 per ml | 0.01~0.04 ml | 0.83 | 3.30 |
| Antiserum- Inaba (2 ml) | 165 | 2 | 82.50 per ml | 0.01~0.04 ml | 0.83 | 3.30 |
| Antiserum - Ogawa (2 ml) | 165 | 2 | 82.50 per ml | 0.01~0.04 ml | 0.83 | 3.30 |
| **Total cost** | | | | | **$5.71** | **$17.08** |

#### Districts excluded from modeling scenarios

**Table S4. Districts excluded from OCV campaign targeting**

These districts were removed from OCV campaign targeting in all model scenarios because they do not encompassing at least one full 5 km * 5 km grid cell, or they overlap completely with a non-residential area (e.g. Lake Rukwa in Tanzania). The incidence rate of these districts might exceed the vaccination threshold under certain scenarios (columns “Bacteriological confirmation capacity” and “Incidence rate threshold”), but they were neither counted as targets nor included in the outcome (e.g. FVP, OCV efficiency, number of targets, etc.) calculation.

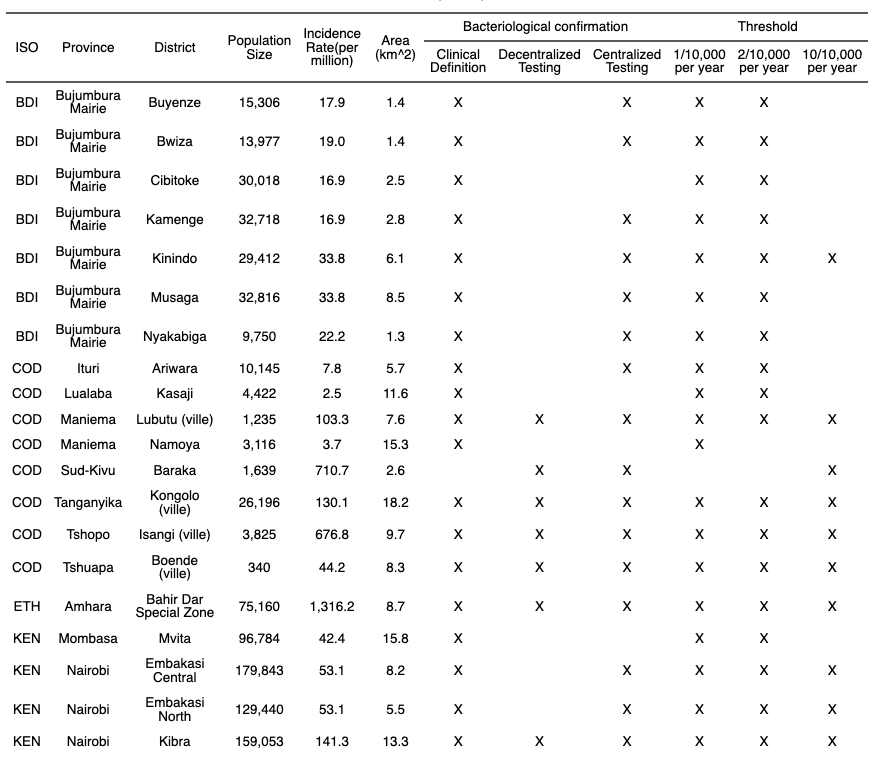

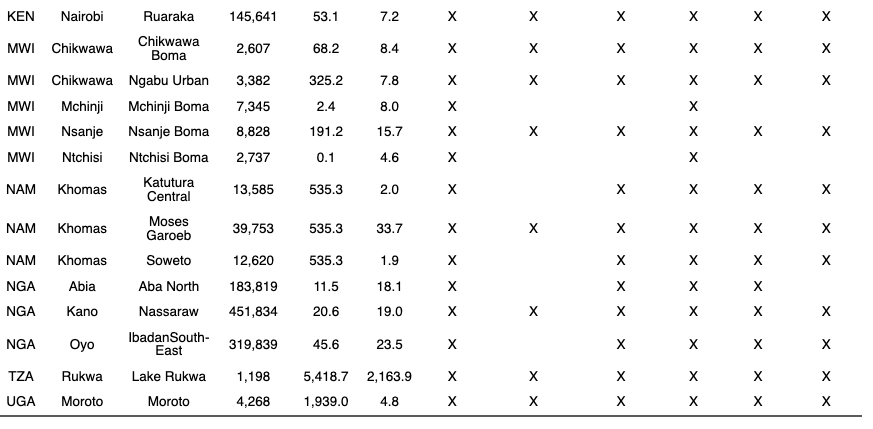

### Supplementary Results

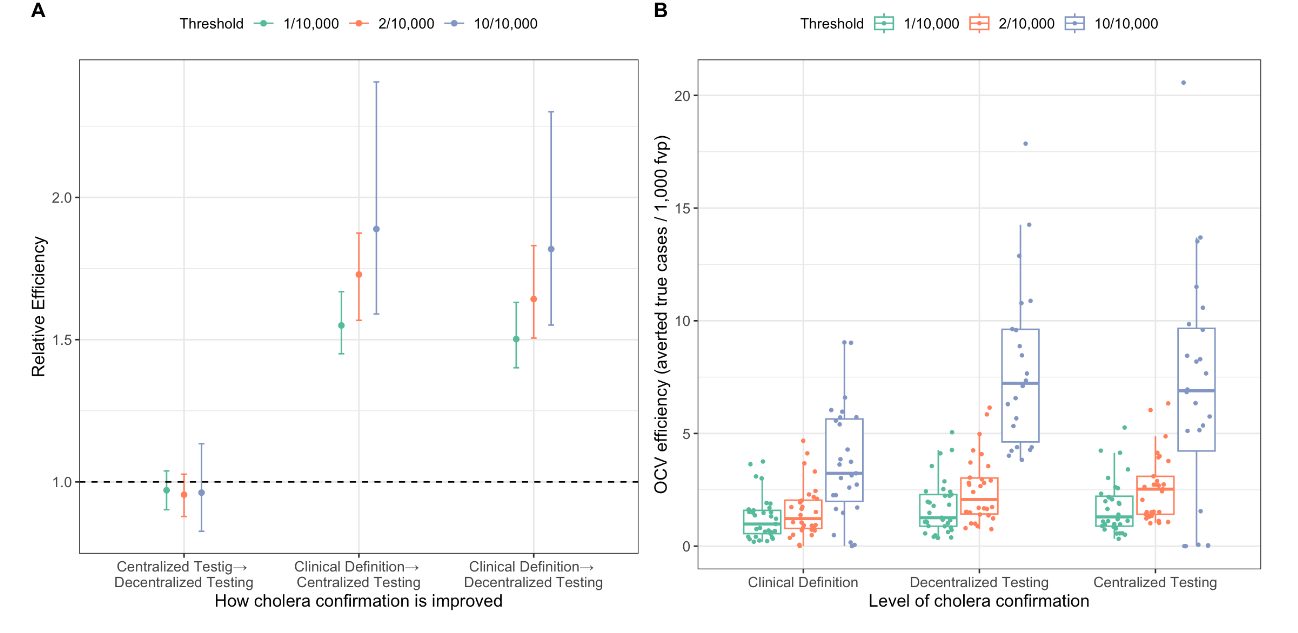

**Figure S1. OCV campaign efficiency by cholera bacteriological confirmation capacity and vaccination threshold for district-level targeting scenarios.** (A) Relative OCV campaign efficiency under different bacteriological confirmation capacity improvements for district-level targeting scenarios. The point represents the median relative efficiency across simulations, and the vertical lines show 95% prediction intervals. The horizontal dashed line represents “No improvement of OCV efficiency” (i.e., relative efficiency = 1). (B) This figure shows the distribution of median OCV campaign efficiency by country. Each point represents the median estimate across simulations for one country, and the boxplot shows the distribution of the country medians.

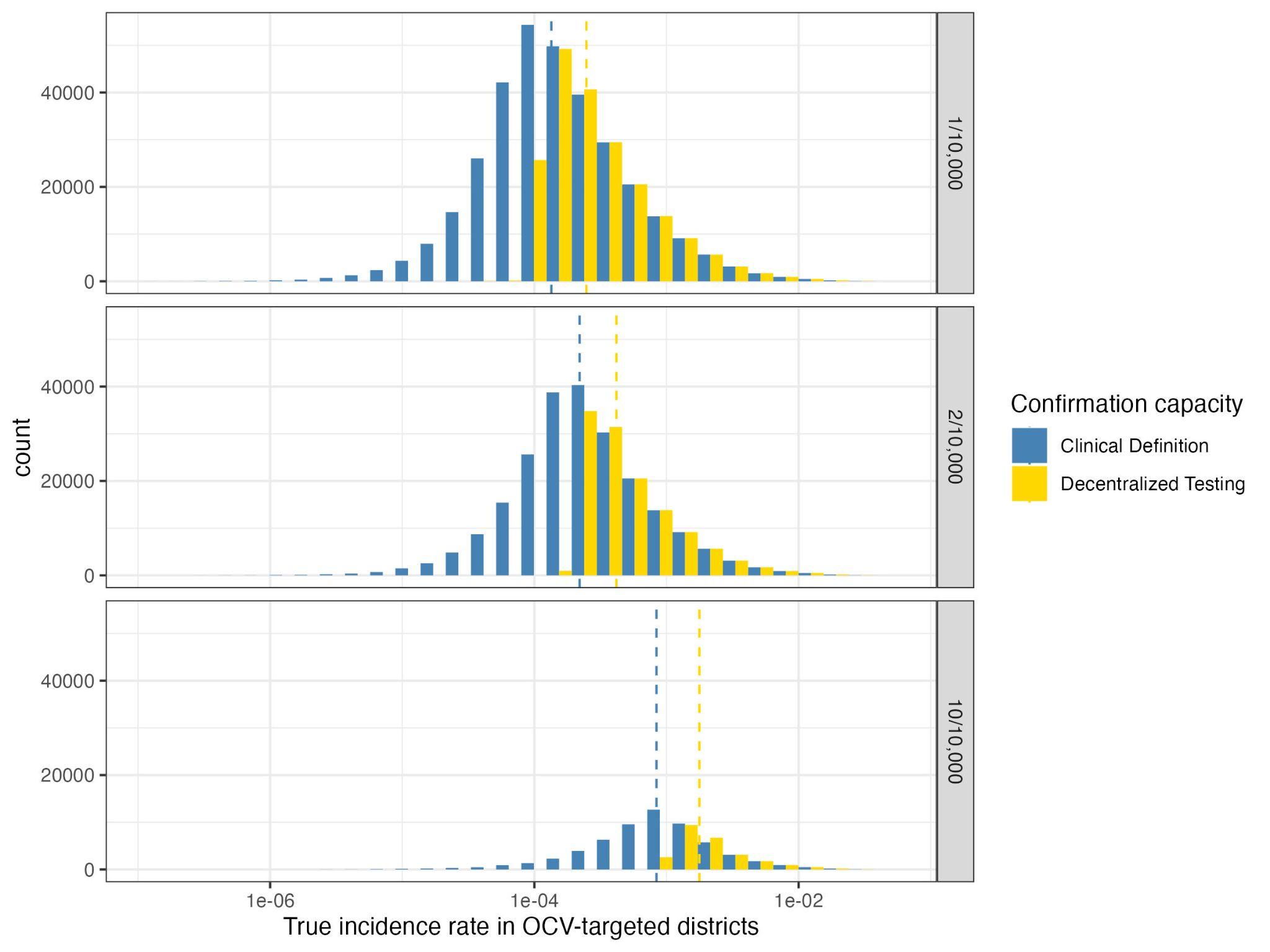

**Figure S2. Baseline true incidence rate of targeted districts.** The histograms show the distributions of true incidence rate of districts targeted by OCV campaigns across 2022-2030 for the three targeting incidence rate thresholds. The incidence rate value is the true incidence rate at the year of being targeted. The steel blue bars represent the distribution of the “Clinical Definition” confirmation scenario, and the gold bars represent the distribution of the “Decentralized Testing” scenario. The incidence rate (x-axis) is displayed on the log scale. The dashed lines mark the medians of the distributions. The medians values for “Clinical Definition” and “Decentralized Testing” are 3.74*10^-4^, 5.78*10^-4^ for the 1/10,000 threshold, 5.16*10^-4^, 8.59*10^-4^ for the 2/10,000 threshold, and 1.38*10^-3^, 2.63*10^-3^ for the 10/10,000 threshold, respectively.

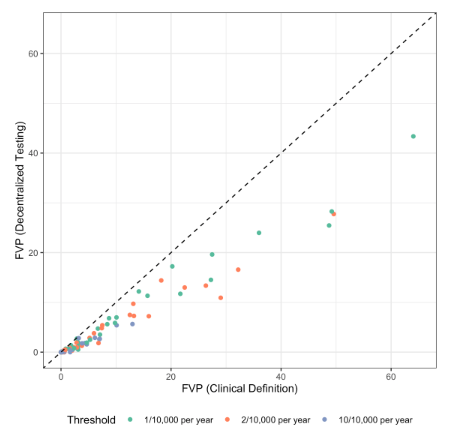

**Figure S3. Country-level fully vaccinated persons (FVP) by introducing “Decentralized Testing” of clinical cholera cases, 2022-2035.** Comparison of fully vaccinated persons (FVP) between “Clinical Definition” and “Decentralized Testing” scenarios, under district-level OCV targeting setting. Each point represents an individual modeled country; the x-axis and y-axis are respectively the median FVPs of the “Clinical Definition” and “Decentralized Testing” scenarios for one specific country; the dashed line marks where y = x.

**Table S5. Complete public health metrics table for all scenarios, 2022-2035.** For all 18 vaccination modeling scenarios, which vary by targeting threshold, campaign targeting scale, and bacteriological confirmation capacity, we report the median estimates and 95% prediction intervals in parentheses for number of fully vaccinated persons (FVP) in millions, true cholera cases averted in millions (averted cases), proportion of true cases got averted in percentage, and true cases averted per 1,000 FVP (OCV efficiency), proportion of FVP living in high incidence rate areas in percentage, total (with repetition) and unique (without repetition) number of administrative units and countries targeted by the OCV campaigns.

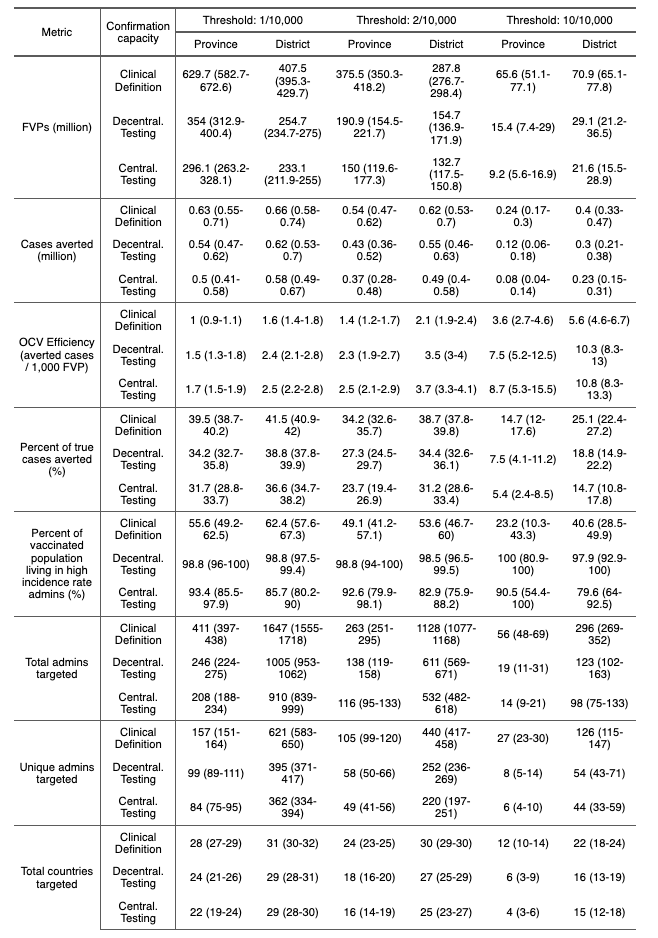
